# High Prevalence and Genotypic Diversity of Persistent *Chlamydia trachomatis* Infections Among South African Adolescent Girls and Young Women: A Tale of Two Cities

**DOI:** 10.1101/2025.01.17.25320646

**Authors:** Smritee Dabee, Shaun Barnabas, Bart Versteeg, Brian Kullin, Shameem Z. Jaumdally, Hoyam Gamieldien, Nonhlanhla Mkhize, Etienne Muller, Venessa Maseko, Katherine Gill, Darren P. Martin, Glenda Gray, Linda-Gail Bekker, David A. Lewis, Heather B. Jaspan, Sylvia M. Bruisten, Jo-Ann S. Passmore

**Affiliations:** Institute of Infectious Disease and Molecular Medicine (IDM), Division of Medical Virology, Dept. Pathology, University of Cape Town, Cape Town, South Africa; DST-NRF CAPRISA Centre of Excellence in HIV Prevention, University of Cape Town, Cape Town, South Africa; Desmond Tutu HIV Centre, University of Cape Town, South Africa; Knowledge institute of the Dutch association of Medical Specialists, Utrecht, Netherlands; Centre for HIV and STIs, National Institute for Communicable Disease, National Health Laboratory Service, South Africa; Perinatal HIV Research Unit, Faculty of Health Sciences, University of the Witwatersrand, Diepkloof, Johannesburg, South Africa; South African Medical Research Council, Cape Town, South Africa; Western Sydney Sexual Health Centre, Western Sydney Local Health District, Parramatta, Australia; Sydney Infectious Diseases Institute & Sydney Medical School-Westmead, University of Sydney, Sydney, Australia; Seattle Children’s Research Institute, University of Washington, Seattle, Washington, USA; Department of Infectious Diseases, Public Health Service Amsterdam, Amsterdam, the Netherlands; Amsterdam Infection and Immunity Institute, Academic Medical Center, University of Amsterdam, Amsterdam, the Netherlands; National Health Laboratory Service, Groote Schuur Hospital, Cape Town, South Africa

**Keywords:** Chlamydia trachomatis, adolescent female, South Africa, molecular epidemiology, multilocus sequence typing, persistence

## Abstract

*Chlamydia trachomatis* (CT) is the most common sexually transmitted bacterial infection globally, significantly affecting adolescent girls and young women (AGYW) in South Africa. This study investigated CT prevalence and genotypic diversity among 298 AGYW from Cape Town and Johannesburg, revealing an overall prevalence of 29.5%. Prevalence was higher in Cape Town (41.6%) compared to Johannesburg (17.4%; p<0.0001). Genetic analysis identified 34 sequence types, including 15 novel variants. Longitudinal data highlighted frequent reinfections or persistent infections despite treatment. These findings underscore the importance of addressing CT’s genetic diversity for improved reproductive health strategies and vaccine development.

**Summary:** *Chlamydia trachomatis* infection is highly prevalent and asymptomatic among South African adolescent girls and young women, with marked regional and genotypic diversity. Frequent persistence or reinfection highlights the need for improved screening, surveillance, and targeted prevention strategies.

## Introduction

Adolescent girls and young women (AGYW) in sub-Saharan Africa are at heightened risk for sexually transmitted infections (STIs), including *Chlamydia trachomatis* (CT), which exacerbates HIV acquisition risks and reproductive complications (1). The asymptomatic nature of most CT infections in women complicates diagnosis and management (2), necessitating a deeper understanding of its epidemiology and genotypic diversity. South Africa shows marked regional variation in CT prevalence and associated complications, potentially reflecting differences in circulating genotypes linked to inflammation, persistence, or increased risk (3–5). Characterising CT genotypes can therefore inform regional epidemiology and support global surveillance efforts.

Here we characterise CT genotypes among AGYW from two urban South African settings (Cape Town and Johannesburg) using high resolution multi-locus sequence typing (MLST) to further explore regional differences in the prevalence of different CT genetic variants (6). These data align with South Africa’s genomic surveillance goals, which could be extremely valuable for monitoring the emergence and spread of pathogenic or treatment-resistant CT strains (7).

MLST typing of CT variants is also useful in that it can (i) indicate whether epidemiologically linked patients experiencing recurrent infections are acquiring them from independent sources, and (ii) differentiate between unrelated new and persistent infections. Inadequately treated CT infections can persist or recur, perpetuating inflammation and increasing the risk of long-term complications.

Evaluating CT genotypes associated with AGYW from two geographically distinct locations in South Africa addresses a gap in our understanding of the interplay between CT genetic diversity and regional epidemiological patterns of CT infections. Closing this gap will inform both general STI and HIV prevention strategies, and the design of CT vaccines tailored to the needs of South African AGYW.

## Methods

### Cohort Description

This study recruited sexually active AGYW aged 16–22 years from two urban centres in South Africa: the Desmond Tutu Health Centre (DTHC) in Cape Town and the Perinatal HIV Research Unit (PHRU) in Johannesburg, using a targeted enrolment strategy with predefined sample sizes (4). These two major urban sites in South Africa, with distinct metropolitan geographies, were selected to allow exploration of shared versus site-specific risk factors. Participants were excluded if they were living with HIV, pregnant, not sexually active, or if they had douched, used spermicides within the past two days, or taken antibiotics within the last two weeks. Ethical approval was obtained from the Universities of Cape Town (UCT HREC #267/2013) and Witwatersrand Research Ethics Committees (WITS HREC #M130745). Written informed consent was provided by participants aged 18 years or older, while those under 18 years provided written assent alongside parental or guardian consent. At enrolment, participants completed a questionnaire capturing socio-demographic characteristics, reproductive health, and sexual behaviour.

### Sample Collection

During each clinical visit, a speculum examination was conducted. A Dacron vulvovaginal swab was collected for STI testing and a lateral vaginal wall FLOQSwab™ sample for preparing a Gram-stained slide for Nugent scoring to diagnose bacterial vaginosis (BV) (4). Vulvovaginal swabs were analyzed for discharge-causing STIs, including *Chlamydia trachomatis* (CT), using real-time multiplex polymerase chain reaction (PCR) assays (4). BV was categorized based on Nugent scoring: scores of 7–10 indicated BV positivity, 4–6 were intermediate, and 0–3 were BV-negative. Participants with positive results received treatment according to South African national guidelines, with a contact slip being given as part of the care protocol (4).

### Chlamydia MLST

The genotypic diversity of *C. trachomatis* was analyzed using MLST and *ompA* genotyping (6)(7). Sequence data from five non-housekeeping gene loci (Uppsala typing scheme) were analysed using the CT database hosted on PubMLST. Allelic combinations that were not present in the CT MLST database were assigned preliminary sequence type (ST) numbers (ST551-ST560). The *ompA* genovars were assigned by comparison to an offline reference database described previously (6). BioEdit was used for sequence editing, with sequences assembled and aligned using the ClustalW tool in MEGA4. Minimum-spanning trees were generated using BioNumerics (Applied Maths, Sint-Martens-Latem, Belgium) to explore genetic relationships between strains. To generate a global CT phylogeny based on the MLST scheme proposed by Klint et al. (8), loci sequences were downloaded for all currently recognised MLST subtypes from the PubMLST database (9), and used to generate in-frame concatenated alignments for each ST using mafft (10), used to construct a maximum likelihood phylogenetic tree using IQ-TREE2 (11) and best fit gene-based partitioned models selected using ModelFinder (12).

### Statistical tests

Differences between sites and age groups were assessed using chi-squared or Fisher’s exact tests for categorical variables and the Kruskal–Wallis test for non-parametric comparisons. All tests were two-sided with p<0.05 considered significant, and analyses were performed using Stata v14.2 (StataCorp, College Station, TX, USA).

## Results

A total of 298 sexually active AGYW from two communities — Cape Town (n=149) and Johannesburg (n=149) — were included in this study. Participants from Cape Town were followed longitudinally over three visits spanning 6–8 months, whereas those from Johannesburg were enrolled for a single visit (Supplementary Table 1). Overall contraceptive use varied significantly between the sites (Kruskal Wallis p<0.0001). While all AGYW in Cape Town used contraception (predominantlyNet-EN; 105/149; 70.5%), in Johannesburg, 90.6% used contraception, with condoms being the most common type (77/149; 63.8%). The prevalence of treatable STIs in the cohort was notably high, affecting 74.2% of participants (221/298). Among these, 10.1% (30/298) had multiple concurrent STIs, and 37.2% (111/298) had both an STI and BV (4).

CT was the most prevalent STI in the study, affecting 88 of 298 participants (29.5%) at enrolment, with prevalence being higher in Cape Town (62/149; 41.6%) than Johannesburg (26/149; 17.4%; p<0.0001). All CT cases were asymptomatic (4). Younger adolescents were disproportionately affected, with a prevalence of 33.8% (69/204) among those aged 16–19 years, compared to 20.2% (19/94) in those aged 20–22 years (Chi-squared p=0.02). Among participants infected with CT, 71.6% (63/88) were co-infected with another STI or had BV (Nugent score 7–10), highlighting the high burden of co-occurring reproductive tract conditions in this population.

*OmpA* typing and MLST were performed on 128 samples with available DNA, including 20/26 cases from Johannesburg and 108/121 from Cape Town. Of these, 18 samples had either insufficient DNA or low bacterial loads. From these CT cases, 34 STs were identified by MLST, with 25/34 circulating in Cape Town and 12/34 in Johannesburg (Figure 1A). Of these, 15 were novel STs not previously reported. The three most common were ST12d (13.3%), ST530b (12%), and ST3 (9.3%). ST12d was the most common ST in Cape Town but was absent in Johannesburg. ST530b was the most common ST in Johannesburg. STs found in Johannesburg but not Cape Town included ST062a, ST086, and ST560.

**Figure 1.**
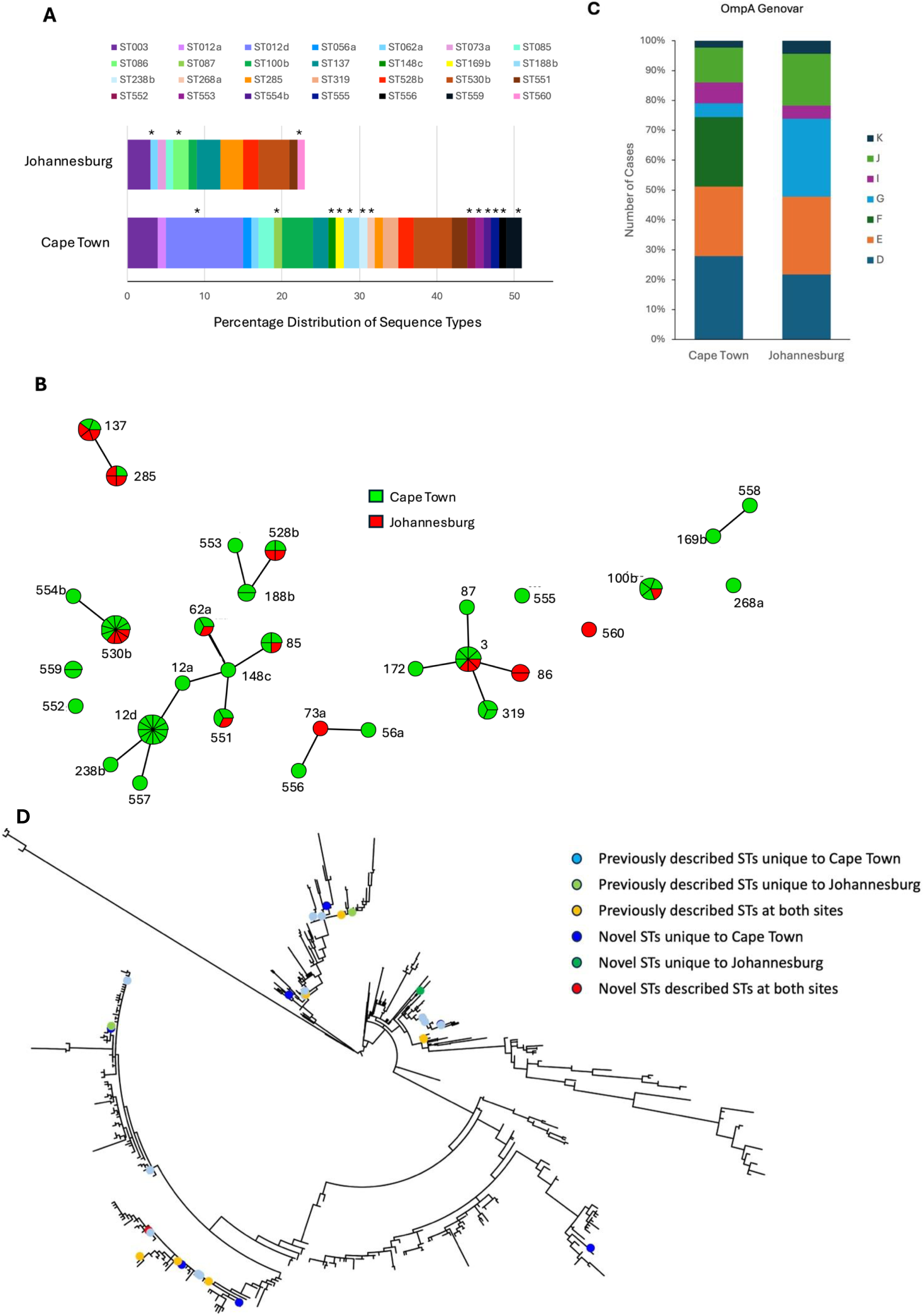
MLST sequence types (ST) and *ompA* genovars in CT isolates from AGYW from Cape Town and Johannesburg. (A) Percentage distribution of MLST STs by region. Each stacked bar represents the proportion of detected STs in Johannesburg and Cape Town. *indicate STs that were unique to each region. (B) Minimum-spanning tree analysis of the identified MLST STs, showing the genetic relationships between STs across regions. The size of the nodes represents the frequency of the sequence type, while connecting lines indicate isolates differing at a single locus. Colours correspond to geographic locations (Cape Town shaded in green and Johannesburg in red). (C) Distribution of *ompA* genovars, stratified by geographic location. (D) Maximum likelihood phylogenetic tree generated from a concatenated gene alignment for all currently recognised CT MLST STs. Coloured circles represent STs detected in the current study.

Despite demographic differences between Cape Town and Johannesburg, a minimum spanning tree analysis revealed no clear clustering of genotypes by location (Figure 1B). Based on sequences from *ompA*, CT genovars from Cape Town and Johannesburg largely overlapped, with genovar E and D being common in both regions (Figure 1C). However, certain notable differences were evident, with genovar F only being found in Cape Town and genovar G being more common in Johannesburg than Cape Town. Mapping STs detected in the current study to the global MLST-based phylogeny (Figure 1D), revealed that there was no clustering by site and that the overall diversity of STs was high.

In Cape Town, where adolescents were followed longitudinally, 33.9% who were infected at visit 1 were also infected with CT at visit 2, and 18.2% were infected at visit 3 (Figure 2A). Among these, 10 individuals tested positive for CT at all three study visits, 10 were infected at the first two visits, and 5 were infected at visits 1 and 3 only. For those infected at >1 study visit, the majority harboured the same ST across visits, suggesting persistent infections or reinfection from the same source (Figure 2B). Limited sample size, particularly after stratification by visit and ST, precludes formal inference about whether prior infection increased the likelihood of infection at follow-up.

**Figure 2.**
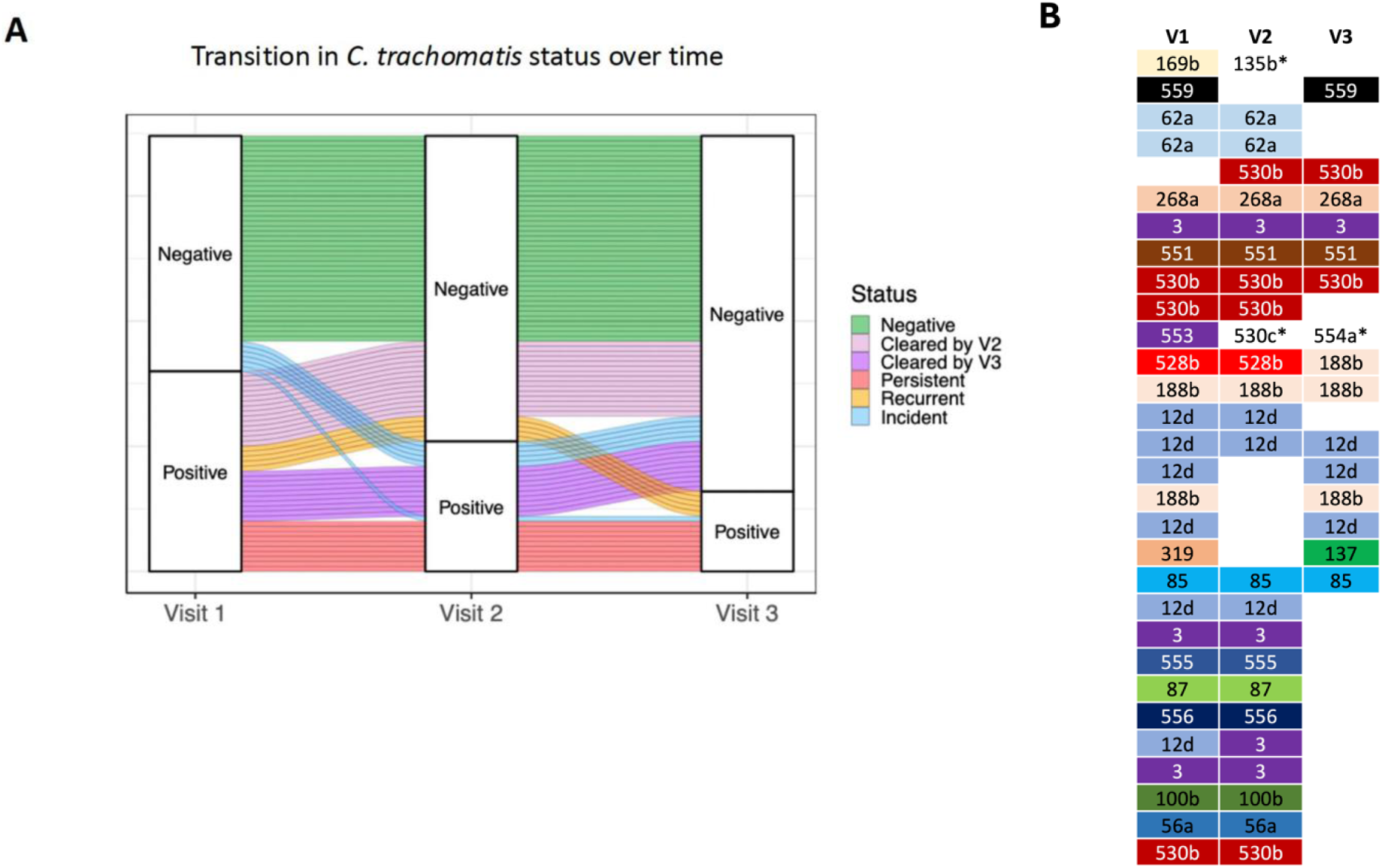
Transition in CT infection status over time in AGYW from Cape Town. (A) Longitudinal changes in CT infection status across three study visits. Participants were categorized based on their infection status at each visit (CT negative [green]; incident [blue], cleared [visit 2: lilac or visit 3: purple], persisted [dark peach] or recurred [light orange]. (B) Comparison of MLST STs identified at multiple visits in AGYW with recurrent infections. The data illustrate whether recurrent infections were associated with the same ST (suggesting persistence or reinfection by untreated partners) or different STs (indicating new infections).

## Discussion

This study highlights the high prevalence, persistence, and genotypic diversity of CT infections among AGYW in South Africa, a group disproportionately affected by STIs and their associated adverse reproductive health outcomes. The findings emphasize the urgent need for targeted interventions to address South Africa’s significant chlamydial burden, particularly its implications for HIV acquisition and complications like pelvic inflammatory disease and infertility. CT prevalence in this cohort was 29.5%, with significant geographic variation: 41.6% in Cape Town compared to 17.4% in Johannesburg. The higher prevalence in Cape Town likely reflects differences in recruitment settings (reproductive health clinics versus general populations) and contraceptive use patterns. However, a limitation of this study is that participants were recruited using a targeted enrolment strategy, and a formal response rate could therefore not be calculated, which may limit the generalisability of the findings. All CT infections were asymptomatic, and younger AGYW (aged 16–19 years) were disproportionately affected, underscoring the need for early sexual health education and routine screening. Co-infections were common, with 71.6% of CT-positive participants also diagnosed with another STI or BV, which may exacerbate genital inflammation and increase HIV susceptibility (13).

Genotypic analysis using MLST revealed significant genetic diversity with 34 STs identified, including 15 novel STs. Regional differences were observed, with ST12d predominant in Cape Town and ST530b in Johannesburg. The lack of geographical clustering indicates widespread circulation of certain genotypes. Notably, whereas globally prevalent *ompA* genovars such as D and E (1) were dominant in both the South African cities studied, another globally dominant genotype, genovar F, was entirely absent in Johannesburg. Genovar G, a less globally common but potentially more pathogenic genotype (14), was detected at a higher frequency in Johannesburg. Given that the infectiousness of CT is significantly influenced by the *ompA* genotype of the infecting strain (15), these findings underscore the importance of regional surveillance to identify locally relevant epidemiological patterns with respect to informing tailored interventions (5). Future studies should investigate the functional implications of these genotypic variations to better understand their role in disease transmission and clinical outcomes.

Persistent or recurrent CT infections were common in the Cape Town cohort, where participants were followed longitudinally. Among those with repeated infections, the same ST was frequently identified across visits, indicating either reinfection from untreated partners or incomplete clearance of infections. This calls attention to the importance of partner notification, treatment, and test of cure to ensure resolution (2) in that persistent infections likely contribute to sustained inflammation, which may exacerbate reproductive health risks, including HIV susceptibility. However, limited sample sizes within STs precluded assessment of type-specific CT persistence or recurrence.

From a public health perspective, addressing the high burden of CT among young women in South Africa requires a multifaceted approach. Although there has been a shift in policy in some countries (like the Netherlands) around the need for ongoing STI screening and treatment, these data from South Africa strongly argue that expanding access to effective and affordable STI POC testing in regions with high HIV burden, integrating partner notification systems, and promoting consistent condom usage are needed for controlling CT infections.

These findings stress the need for vaccines targeting CT, particularly those tailored to the most prevalent genotypes in high-burden regions (8). Collar and colleagues (1) present compelling evidence emphasizing the necessity for CT vaccine strategies to account for regional variations in CT genotypes. This approach is critical to ensuring that interventions are not only effective but also equitable, addressing the unique epidemiological and genetic diversity of CT across diverse communities. Genomic surveillance is set to play a pivotal role in addressing CT burden (5). Incorporating genetic diversity data into vaccine development pipelines should also ensure broader protection against locally prevalent rather than just globally dominant CT genotypes (5).

In conclusion, this study provides an analysis of CT prevalence, persistence, and genotypic diversity among AGYW in South Africa. The findings emphasize the need for targeted interventions to mitigate the health risks associated with CT. By integrating genomic surveillance, public health initiatives, and community-based strategies, we can address the multifaceted burden of CT and improve reproductive health outcomes for AGYW in South Africa.

## Data Availability

All data produced in the present study are available upon reasonable request to the authors

## Acknowledgements

We extend our gratitude to the WISH study team, with special thanks to Ms. Penelope Ngcobo for her efforts in participant recruitment and Sr. Janine Nixon. We are deeply appreciative of the young women who participated in the study, without whom this research would not have been possible. The data underlying this article are available in the PubMLST Chlamydia trachomatis database at https://pubmlst.org/organisms/chlamydia-trachomatis, and can be accessed using the assigned multilocus ST identifiers, including novel STs reported in this study.

