## Supplementary Table 1 for "High Prevalence and Genotypic Diversity of Persistent *Chlamydia trachomatis* Infections Among South African Adolescent Girls and Young Women: A Tale of Two Cities"

**Supplementary Table 1.** Characteristics for study cohort

| Characteristics | All  (n=298) | Johannesburg  (n=149) | Cape Town  Visit 1 (n=149) | Cape Town  Visit 2 (n=128) | Cape Town  Visit 3 (n=93) | p-value* |
| --- | --- | --- | --- | --- | --- | --- |
| Age | 18 (17-20) | 18 (17-20) | 18 (17-20) | - | - | ns |
| Age of sexual debut | 16 (15-17) | 16 (16-17) | 16 (15-17) | - | - | ns |
| Lifetime # partners | 2 (1-13) | 2 (1-10) | 2 (1-13) | - | - | ns |
| Condoms (always) | 69/267 (25.8%) | 49/138 (36%) | 20/129 (16%) | - | - | 0.0002 |
| Condoms (last sex) | 190/267 (71.2%) | 94/138 (68%) | 96/129 (74%) | - | - | ns |
| Sex w HIV+ partner | 20/267 (7.5%) | 7/138 (5%) | 13/129 (10%) | - | - | ns |
| Transactional sex | 1/267 (0.4%) | 0/138 (0%) | 1/129 (1%) | - | - | ns |
| Contraceptives  Ever pregnant | 284/298 (95.3%)  64/267 (24%) | 135/149 (90.6%)  30/138 (22%) | 149/149 (100%)  34/129 (26%) | - | - | <0.0001  ns |
| Anal sex (ever) | 11/267 (4.1%) | 3/139 (2%) | 8/129 (6%) | - | - | ns |
| STI frequency: |  |  |  |  |  |  |
| Any STI | 221/298 (74.2%) | 101/149 (67.8%) | 120/149 (80.5%) | 97/128 (75.8%) | 70/93 (75.3%) | 0.02 |
| *C. trachomatis* | 88/298 (29.5%) | 26/149 (17.4%) | 62/149 (41.6%) | 43/127 (33.9%) | 16/88 (18.2%) | <0.0001 |
| *N. gonorrhoea* | 24/298 (8.1%) | 7/149 (4.7%) | 17/149 (11.4%) | 8/127 (6.3%) | 7/88 (7.9%) | 0.05 |
| *T. vaginalis* | 16/298 (5.4%) | 5/149 (3.4%) | 11/149 (7.4%) | 10/127 (7.9%) | 6/88 (6.8%) | ns |
| *M. genitalium* | 11/298 (3.7%) | 5/149 (3.4%) | 6/149 (4.0%) | 7/127 (5.5%) | 6/88 (6.8%) | ns |
| Multiple STIs | 30/298 (10.1%) | 6/143 (4.0%) | 24/149 (16.1%) | 44/128 (34.4%) | 23/88 (26.1%) | 0.001 |
| BV (Nugent 7-10) | 135/292 (46.2%) | 64/144 (44.4%) | 71/148 (48.0%) | 62/126 (49.2%) | 39/84 (46.4%) | ns |
| Any STIs+BV | 111/298 (37.2%) | 51/149 (34.2%) | 60/149 (40.3%) | 46/128 (35.9%) | 31/93 (33.3%) | ns |

*Fisher’s Exact test
